# Identifying stable-against-mutations viral epitopes in SARS-CoV-2

**DOI:** 10.1101/2022.10.13.22280980

**Authors:** Ildefonso M. De la Fuente, Iker Malaina, Maria Fedetz, Maksymilian Chruszcz, Gontzal Grandes, Oleg Targoni, Antonio Abel Lozano-Pérez, Eyal Shteyer, Ami Ben Ya’acov, Agustín Gómez de la Cámara, Alberto M. Borobia, Jose Carrasco-Pujante, Jose Ignacio Pijoan, Carlos Bringas, Gorka Pérez-Yarza, Alberto Ouro, Ivan Sanchez, Jesus M. Cortes, Michael J. Crawford, Varda Shoshan-Barmatz, Vladimir Zhurov, José I. López, Shira Knafo, Magdalena Tary-Lehmann, Toni Gabaldón, Miodrag Grbic

**Author notes:** **Corresponding Authors:** -Ildefonso M. De la Fuente, Instituto CEBAS-CSIC., Campus Universitario de Espinardo., Espinardo., 30100 Murcia. ESPAÑA., -Iker Malaina Celada, UPV/EHU, Faculty of Science and Technology, Department of Mathematics, Barrio Sarriena, s/n, 48940 Leioa, Spain. Equal contribution (Ildefonso M. De la Fuente, Iker Malaina).

## Abstract

We have developed a computational method “Multi-Stable Epitope Sequencer” to predict mutation-resistant regions with stability against future viral variability. At the beginning of the pandemic, this approach allowed us to identify a set of eight SARS-CoV-2 spike protein sequences that had the potential to be mutationally stable. We have tested this methodology on the SARS-CoV-2 viral linages that occurred throughout the COVID-19 pandemic. These eight peptide sequences (epitopes) have been preserved in 97% of all SARS-CoV-2 lineages reported in the CoV-GLUE dataset during the pandemic period. Likewise, more than 90% of these peptides remained invariable across the 49 predominant viral variants circulating throughout the pandemic (ECDC-WHO). In addition, the eight selected peptides were preserved in 94.1% of all 28 variants considered of most interest in the CoV-GLUE project. Our analyses confirm the predicted mutational stability of the eight selected short viral peptides over the entire COVID-19 pandemic.

## 1. Introduction

The greatest challenge in the development of successful preventive vaccines lies in identifying antigenic epitopes likely to be resistant to future mutations, and capable of inducing an effective, safe, and long-lasting immune response^1^. The enormous difficulty of accommodating the inevitable and complex mutational dynamics of viruses has hampered the development of efficient vaccines.

Different research groups are trying to develop mathematical-computational tools that predict likely mutational hotspots so that they can be avoided. For example, Maher et al.^2^, and Obermeyer et al.^3^ have proposed approaches that rely upon large extant sequence data sets to predict the amino acids most likely to change in SARS-CoV-2. In this line of work, Meynard-Piganeau et al. have applied neural networks to optimize the prediction of MHC molecules and T cell receptors in peptides^4^. Some initiatives have attempted to overcome these challenges by designing strategies to predict or retrospectively identify conserved, variant-resistant neutralizing epitopes^5-11^.

Tools from computational biology have been also used for a variety of objectives in fighting COVID-19, such as, for instance, optimizing antibodies^12^ identifying inhibitors of the interaction of the Spike protein with ACE2 receptor^13^, modeling^14^, designing protein inhibitors^15^, obtaining epitope maps^16^ or designing candidate vaccines^17-19^. “In silico” prediction of the neutralizing antibody response^20^, and combinatorial optimization to select vaccine epitopes on the basis of the 22 known variants of concern of SARS-Cov-2^21^ have been also carried out. The computational design of vaccines is developing increasingly, thanks to the constant improvement of mathematical-bioinformatics tools, and its results after adequate tests will help advance this important area of research^20, 22-31^.

Over the last 11 years, we have developed a methodology that allows the computational design-selection of specific epitopes with high probability to be stable against future mutations over a long period of evolution. In contrast to the classical methods intending to protect against an individual variant of the virus, our methodological platform, called “Multi-Stable Epitope Sequencer” identifies peptides with immunogenic capacity and affinity to the HLA Class I and Class II histocompatibility antigens. The method also takes into account the absolute frequency of the different HLA alleles present in the world population. Thus, our approach would favor the selection of peptides with higher affinity to the most prevalent histocompatibility molecules. In addition, these potential epitopes are essentially characterized by a higher probability of remaining stable over a determinate viral evolution. Our approach, based upon combinatorial mathematics, advanced computational techniques, and immune-informatics tools, provides a new methodology to obtain candidate epitopes most likely to remain stable against future mutations for long periods of time (for more details see “Methods” section 5.1-5.3).

We have tested this methodology on the SARS-CoV-2 viral lineages. At the beginning of the COVID-19 pandemic (February-March 2020), when only 22 viral genomes deposited in the GISAID project could be analyzed, we identified a set of eight short SARS-Cov-2 Spike protein sequences (patent register #P202030467, Spain, May 2020) using our predictive approach. These peptides (epitopes) exhibited the potential to be mutationally stable.

We here document the degree to which these eight epitopes remained stable throughout the entire period of the COVID-19 pandemic. The World Health Organization had declared a Public Health Emergency of International Concern (PHEIC) for SARS-CoV-2 infection on January 30, 2020, which represents its highest level of alarm under international health regulations. This institution declared the outbreak of COVID-19 a pandemic on March 11, 2020. Finally, WHO officially declared the end of the pandemic phase on May 5, 2023. Our aim is to confirm the predicted mutational stability of the eight selected short viral peptides over the period (March 11, 2020 to January, 2024).

Specifically, we conducted a stability study of the selected epitopes using Spike mutations reported in the CoV-GLUE dataset (the Pango Lineage Resources)^32^ for the all 1,514 SARS-CoV-2 lineages originated during the pandemic period, showing that our epitopes are conserved in at least 97% of them. In addition, we analysed the mutational stability of the eight-epitope pool in the 49 predominant SARS-CoV-2 variants considered by ECDC-WHO, taking into account Spike-defining mutations listed by the GISAID-Outbreak Project^33^, and the sequence stability of the eight-epitope pool was 90.9% in the variants considered by ECDC-WHO. Likewise, we have observed that the eight selected peptides have been preserved in 94.1% of 28 variants considered of most interest in the CoV-GLUE project.

We finally analyzed 3,362 complete viral genomes from the Nextstrain project^34^ and 5,228,435 sequences, including mutations with low frequencies (0.0001) catalogued by CoV-GLUE database^35^, and the obtained results have shown that our epitopes are located in Spike protein cold spots, conserved regions with very low mutational frequency. The obtained results have shown that all our epitopes are located in Spike protein regions with very low mutational frequency. In addition, the peptide mapping analysis shows that the selected epitopes are exposed to solvents in the S-spike protein structure; therefore, none of these epitopes are completely buried into the Spike protein trimer, which allows accessibility to antibodies.

In summary, we designed eight short epitopes with the potential to be resistant to mutations when the pandemic started; this study confirms the mutational stability of epitopes selected by our methodology to anticipate possible future variations.

These results show that the “Multi-Stable Epitope Sequencer” computational method is able to predict epitopes with SARS-CoV-2 which are likely to remain stable for long periods of time. This approach opens up a perspective for new methodological procedures able to serve as a basis for the development of universal vaccines, which pre-emptively forestall most or all future variants, and that provide neutralizing antibody therapies by selecting stable epitopes against future mutations.

## 2. Results

### 2.1. Selection of eight SARS-CoV-2 mutationally stable epitopes at the beginning of the COVID-19 pandemic

We first quantitatively analyzed the characteristics of the viral Spike protein using our computational-predictive method, “Multi-Stable Epitope Sequencer” (see “Methods” section 5.1-5.3), in order to identify a set of SARS-CoV-2 sequences (epitopes) that had the potential to be mutationally stable.

The analysis identified a combination of short peptides (P1-P8) with sequences comprising 10 to 24 amino acids with robust stability against possible pathogen mutations. This epitope pool is also characterized by high immunogenicity and affinity to the HLA antigens, covering 85% and 100% of the most frequent Class I and Class II alleles respectively. The location of these peptides within the Spike protein, as well as their corresponding amino acid sequences, is shown (Fig. 1). Although the algorithm scanned all Spike regions without bias, the eight short peptides (P1-P8) identified were mainly localized to domains critical for viral entry into the host cell, including two peptides located near the angiotensin-converting enzyme 2 (ACE2) binding site (Fig. 1).

**Fig. 1.**
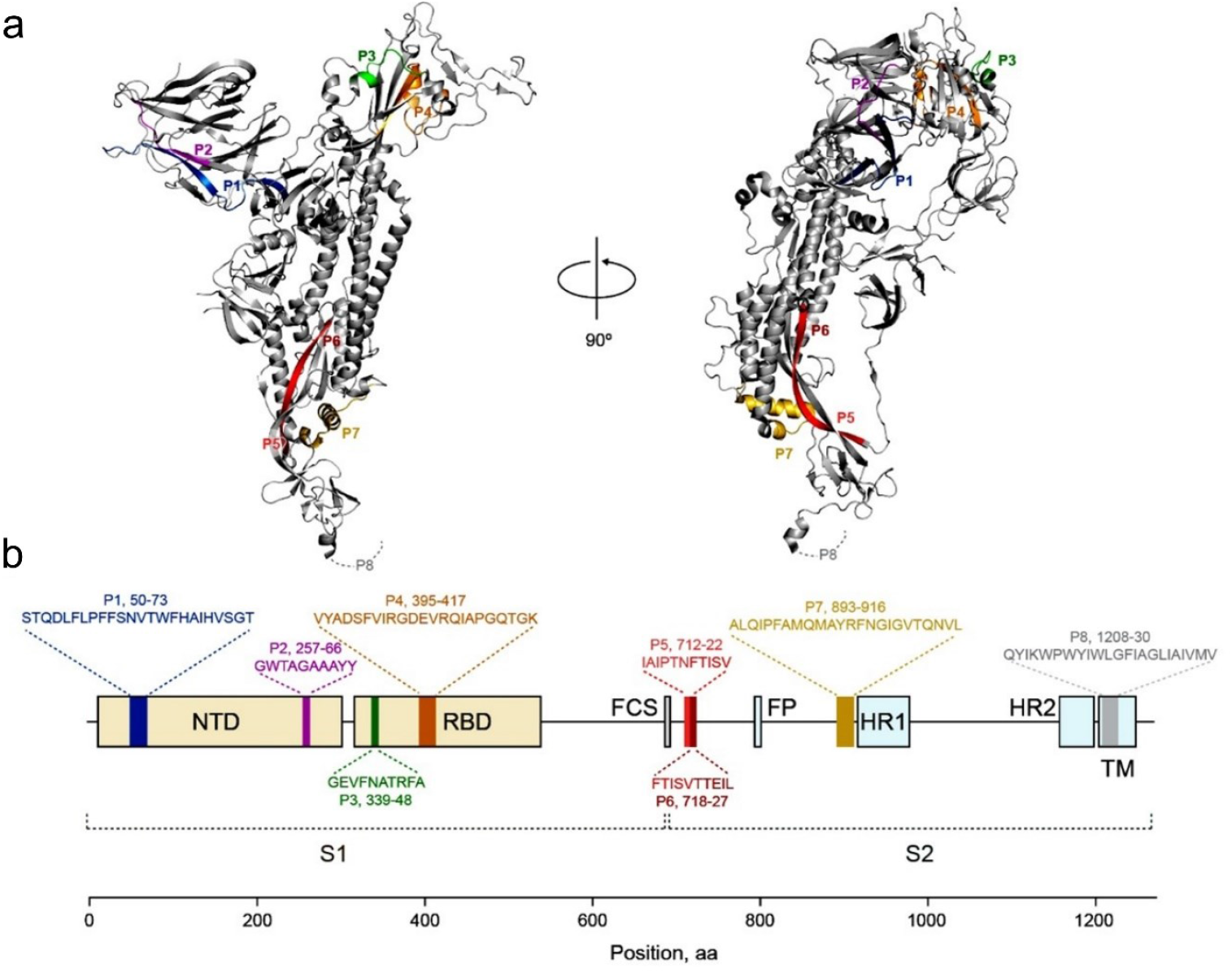
Peptide sequences and location of the eight regions stable against mutations identified at the beginning of the COVID-19 pandemic. **a** Structure of the Spike protein and locations of the eight identified peptides. **b** The complete sequences of the eight epitopes and their respective locations, where P3 and P4 peptides are located in the RBD (Receptor Binding Domain). Noteworthily, our methodology studied the S-structure as a whole, without preselecting any determinate region. S1 – Receptor Binding Domain; S2 – Membrane Fusion Domain; NTD – N-Terminal Domain; FCS – Furin Cleavage Site; FP – Fusion Peptide; HR1 and 2 - Heptapeptide Repeat Sequence; TM – Transmembrane domain.

Our epitope selection was developed by February-March 2020 when only 22 entire viral Spike protein genomes had been deposited in the GISAID project database^36^. The sequences of the eight peptides (P1-P8) were submitted to a formal registration procedure (May 2020, patent register #P202030467, Spain).

### 2.2 Stability of the selected epitopes (P1-P8) against non-synonymous Spike mutations in all 28 variants of most interest in CoV-GLUE dataset

First, we assessed the stability of the selected epitopes against Spike non-synonymous mutations in the all 28 variants of most interest reported in the CoV-GLUE dataset (Fig. 2) throughout the entire period of the COVID-19 pandemic (March, 2020 to December, 2024). The results indicated that the peptides (P1-P8) exhibit a very low average probability of mutation. Specifically, the P1 epitope is conserved in 95% of the lineages; the P2 epitope in 99.8%; the P3 epitope in 89.1%; the P4 epitope in 89.2%; the overlapping the P5-6 epitope, in 96.1%; epitope P7 in 98.2%; and finally, the P8 epitope in 91.6% of the lineages. This translates to an overall conservation rate of 94.1%, with a confidence interval *CI*=(90.19%,98.08%).

**Fig. 2.**
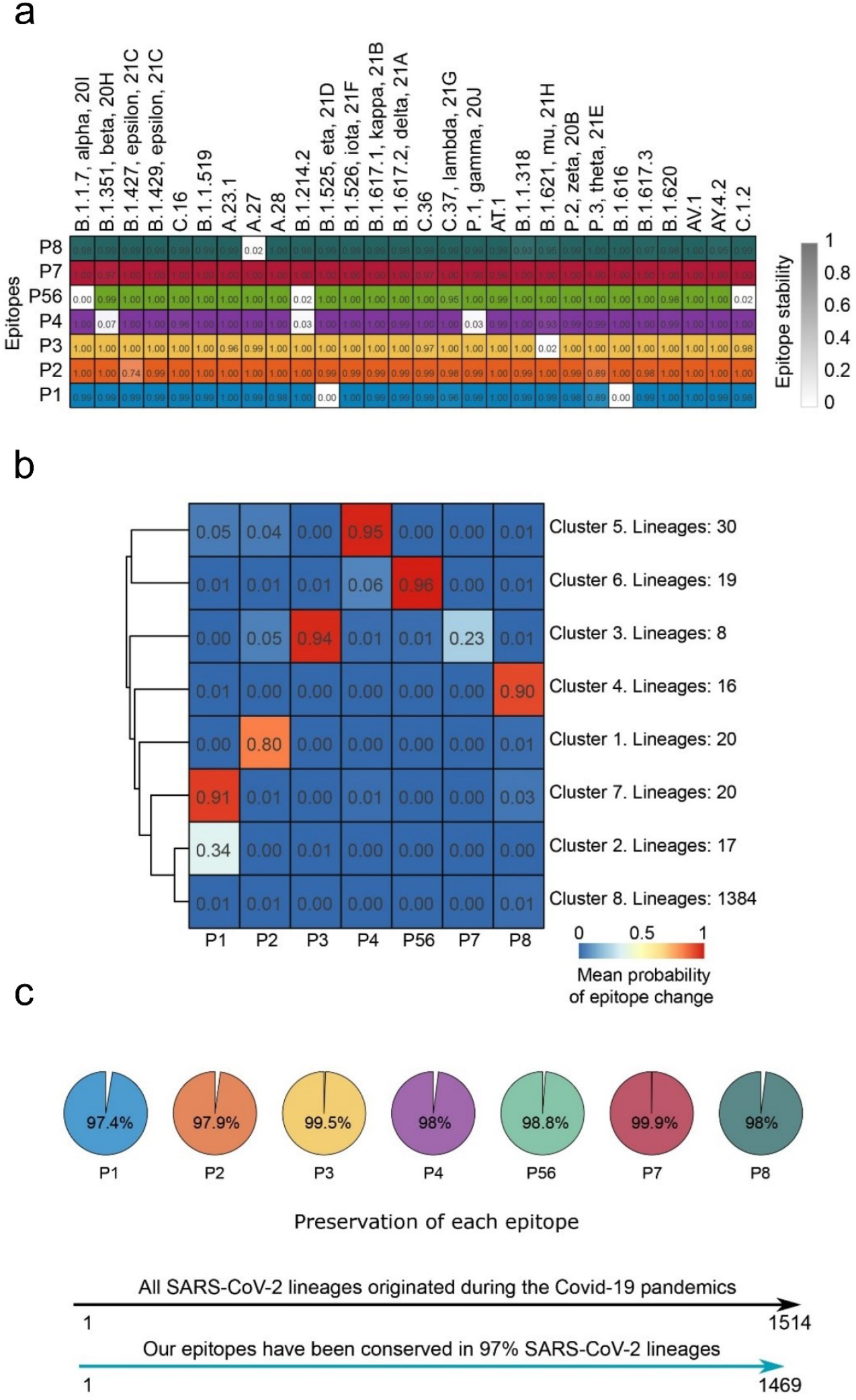
Stability of the selected epitopes against all non-synonymous mutations in the 28 lineages considered of most interest, and in 1514 lineages originated during the Covid-19 pandemic. **a** Stability of the selected epitope regions against all Spike mutations in the CoV-GLUE dataset in the 28 lineages of most interest. **b** k-means clustering of probabilities of observation of non-synonymous mutation within regions of selected epitopes across 1514 lineages in the CoV-GLUE dataset. The distance is Euclidean, the linkage is Ward’s, k = 8. Our selected epitopes are conserved in 97% of lineages. **c** The percentage of conservation of the epitopes (P1-P8).

### 2.3 Stability of the selected P1-P8 peptides in all the 1514 known SARS-CoV-2 lineages curated in CoV-GLUE dataset

We next studied the genetic stability of the P1-P8 peptides surveyed the non-synonymous mutations reported in the CoV-GLUE dataset of all 1,514 known SARS-CoV-2 lineages documented during the pandemic. The k-means clustering analysis of mutation probabilities of all lineages (Fig. 2b) indicates that the vast majority (1384 out of 1514, or 91.4%; cluster 8) exhibit an extremely low average probability of mutation within the selected epitopes (P1-P8). Among the rest of the lineages (130), 85 exhibit low mutational probability, while only 45 present a high probability of change, limited to a specific region of a single epitope. Our results indicate that the P1 epitope was stable in 97.4% of the lineages; the P2 epitope, 97.9%; the P3 epitope, 99.5%; the P4 epitope, 98%; the overlapping the P5-6 epitope, 98.8%; the P7 epitope, 99.9%; and, finally, the P8 epitope was stable in 98% of the lineages. The percentage of conservation of all short peptides (P1-P8) indicates an overall preservation ratio of 98.5%, with a confidence interval *CI*=(97.65%, 99.34%). Our data demonstrate, with 95% of confidence, that the selected epitopes (P1-P8) have been conserved in at least 97% of SARS-CoV-2 lineages that arose during the Covid-19 pandemic

### 2.4. Stability of the eight selected epitopes (P1-P8) among the main SARS-CoV-2 variants reported by ECDC-WHO

The evolution of the SARS-CoV-2 genome has generated a complex cascade of lineages derived from a common ancestor (Fig. 3a). We have analyzed the 49 SARS-CoV-2 predominant viral variants circulating throughout the pandemic to evaluate the mutational stability of the eight selected peptides (P1-P8), taking into account the defining Spike mutations (GISAID-Outbreak project).

**Fig. 3.**
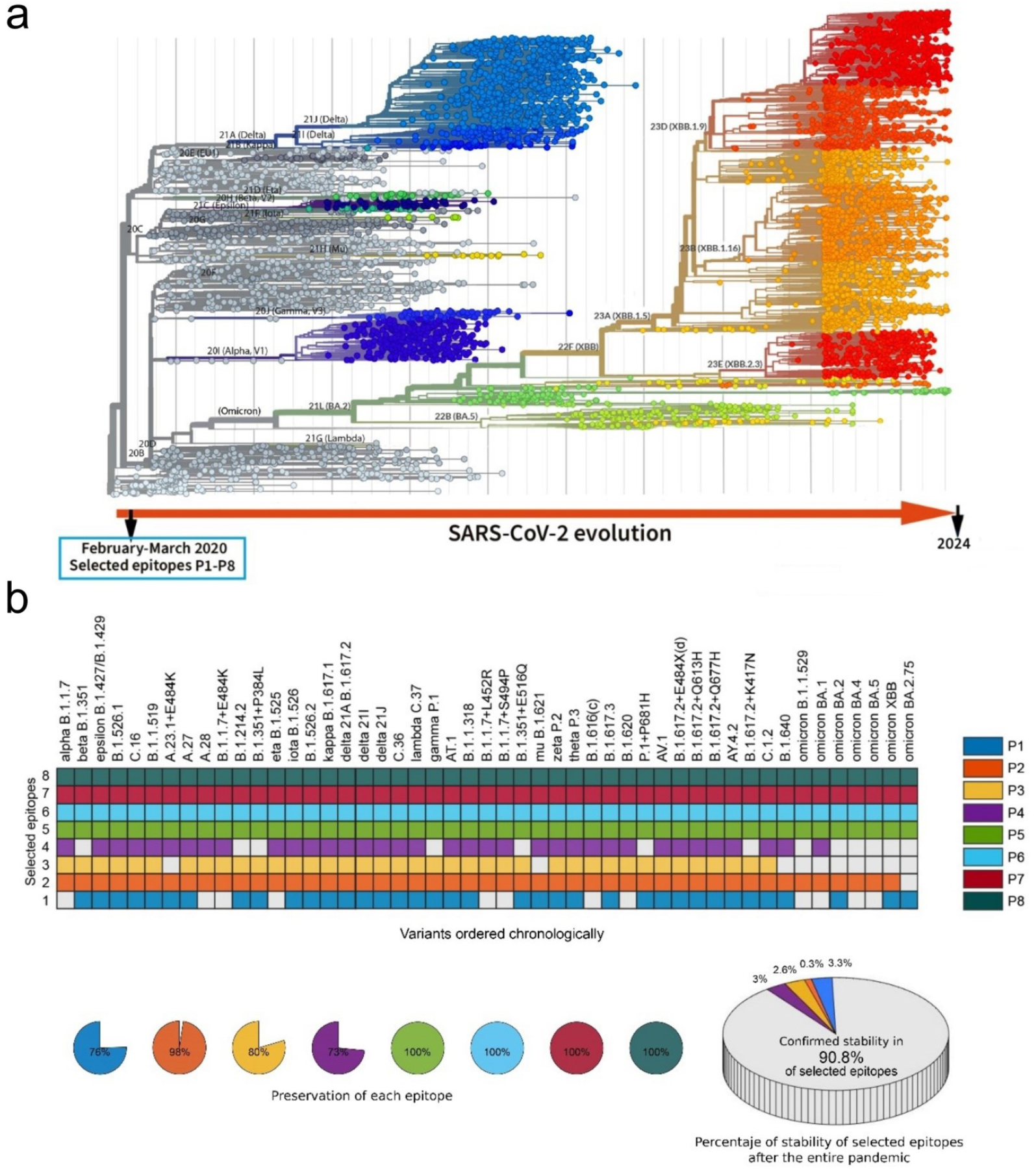
Identified epitope stability against defining mutations in the main 49 SARS-CoV-2 variants. **a** Phylogeny summary of the SARS-CoV-2 virus throughout the entire pandemic the period (March 2020 to January 2024). **b** The panels illustrate the mutational stability of the selected epitopes in each variant of most interest. The percentage of stability of the selected eight peptides (P1-P8) in each of the 49 variants considered (ECDC-WHO) is 90.82% in total, with a confidence interval of *CI*=(87.55%, 93.29%).

Fig.3a shows the phylogeny of the SARS-CoV-2 virus until January 2024 (nextstrain.org)^37^. The origin of the mutational cascade was established in the first viral samples obtained in China at the beginning of the pandemic^38^. The 49 SARS-CoV-2 variants *of most interest* (as determined by the European Centre for Disease Prevention and Control - ECDC) are shown (Fig. 3b). The list includes variants of concern (VOC), Variants Under Monitoring (VUM), and De-escalated variants (ecdc.europa.eu)^39^. The ECDC traces a wider number of variants than those the considered by the World Health Organization (who.int)^40^. The variants defined by the ECDC exhibit “significant potential for transmissibility, severity, and/or immunity likely to have an impact on the epidemiological situation based on properties analyzed by combined genomic, epidemiological, and in vitro pieces of evidence” (ecdc.europa.eu). All the main variants of most interest were studied. Specifically, the total number of variants considered in our study (n=49), the corresponding Pango lineage, WHO-ECDC nomenclature, and the percentages of stability for the eight epitopes in each variant, are provided (Fig. 3b). All the characteristic mutations for the analyzed lineages (non-synonymous substitutions or deletions that occur in > 75% of sequences within each variant) were obtained from the GISAID-Outbreak project^41^.

The study of mutations in each selected peptide shows (Fig. 3b, down) that the P5, P6, P7, and P8 epitopes never exhibit mutational modification. The P4 epitope appears mutated in thirteen variants out of 49, displaying a mutational probability of 0.265, with a confidence interval of *CI*=(0.162,0.403). The P3 epitope is mutated in ten variants with a mutational probability of 0.204 and a confidence interval of *CI*=(0.115, 0.336). The P2 epitope appears mutated in a single variant, displaying a mutational probability of 0.02, with a confidence interval of *CI*=(0.004,0.107). Finally, the P1 epitope is mutated in twelve variants and exhibits a mutational probability of 0.244 with a confidence interval of *CI*=(0.146, 0.381).

We designed eight short peptides with the potential to be stable against mutations when the pandemic started and now, this analysis confirms that the stability percentage of the selected eight-epitope pool (P1-P8) in the 49 SARS-CoV-2 variants of most interest (ECDC-WHO) is 90.82% with a confidence interval of *CI*=(87.55%, 93.29%).

Therefore, we observe a strong agreement in the subset of 28 variants (taking into account non-synonymous mutations reported by CoV-GLUE dataset, Fig. 2a) with the analysis focused on defining mutations of SARS-CoV-2 variants catalogued by ECDC (GISAID-Outbreak project) (Fig. 3).

### 2.5. Stability against mutations of the eight epitopes (P1-P8) against 3,362 complete genomes and 5,228,435 SARS-CoV-2 sequences

Finally, we conducted a stability study of the selected epitopes (P1-P8) using the public data at GISAID^36^, which included 3,362 complete genomes across 1,514 SARS-CoV-2 lineages (available at Nextstrain)^34^. We used this dataset to calculate the entropy per site and the number of mutational events across the strain tree in codons within and outside our selected peptides. In addition, we measured protein variation using GISAID hCoV-19 sequences available at the CoV-GLUE database^35^, which includes very rare mutations (0.0001).

An analysis covering the distribution of non-synonymous mutation frequencies, site entropies, and a number of inferred mutational events per site across the SARS-CoV-2 phylogeny is shown (Fig. 4a–d). This study (see “Methods” section5.6) reveals that the non-synonymous mutations are not uniformly distributed over the SARS-Cov-2 S-protein; consequently, we can observe regions with high or low mutation frequency. Specifically, all selected epitopes (P1-P8) are located in the S-spike protein cold spots (conserved regions with very low mutational frequency).

**Fig. 4.**
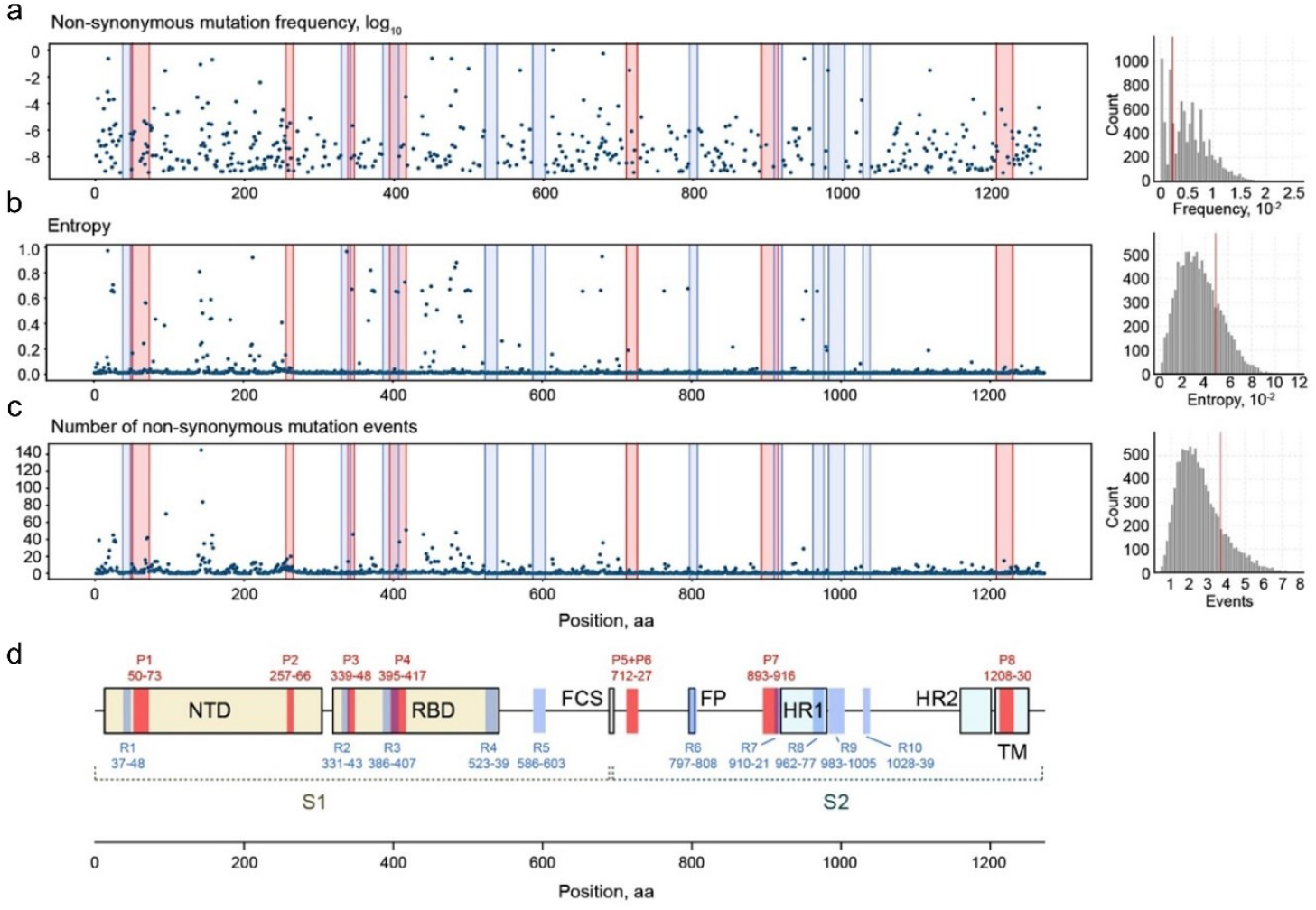
Stable SARS-CoV-2 S-spike sequences against mutations. **a** Distribution of non-synonymous mutation frequencies. **b** Site entropies, normalised Shannon entropy where 1 is maximal entropy. **c** A number of inferred mutational events per site across the SARS-CoV-2 phylogeny are shown as dot plots for all residues of the S-spike protein. **d** Schematics of S-protein selected peptides, overlying data in (**c**). Bar plots on the right show the distribution of average values for 10,000 randomly chosen combinations of epitopes of similar size. Vertical red bars indicate the value for our combination of eight epitopes (P1-P8): frequency of 0.0023 mutations per site, 3.73 mutational events per site, and relative entropy of 0.049. Vertical blue bars correspond to the set of ten highly conserved peptide sequences, detected but not selected (R1-R10), see Methods section 5.4.

In this post-hoc sequence analysis, we found another ten short peptides with low mutation rate (Fig. 4a–d, marked in blue R1-R10). The complete sequence of these peptides (R1-R10, detected but not selected), is indicated in the “Methods” section. In total, all 18 detected mutation-stable regions have an average amino-acid length of 16.22±5.54 (mean ± SD), with a confidence interval *CI*=(13.47, 18.98). Note that our predictive method selected eight short SARS-CoV-2 S-spike peptides (P1-P8), among millions of combinations of possible amino acid sequences including the previously mentioned ten highly conserved peptide sequences (Fig. 4). This latter set of stable Spike protein peptides (R1-R10) was not finally chosen by our computational approach because analytical results excluded them on the basis of lower potential to induce an immune response (see “Methods” section5.4).

### 2.6. Peptide mapping and solvent expositions of eight selected epitopes stable against mutations

We mapped the selected epitopes (P1-P8) on the three-dimensional structure of SARS-CoV-2 Spike protein (Fig. 5).

**Fig. 5.**
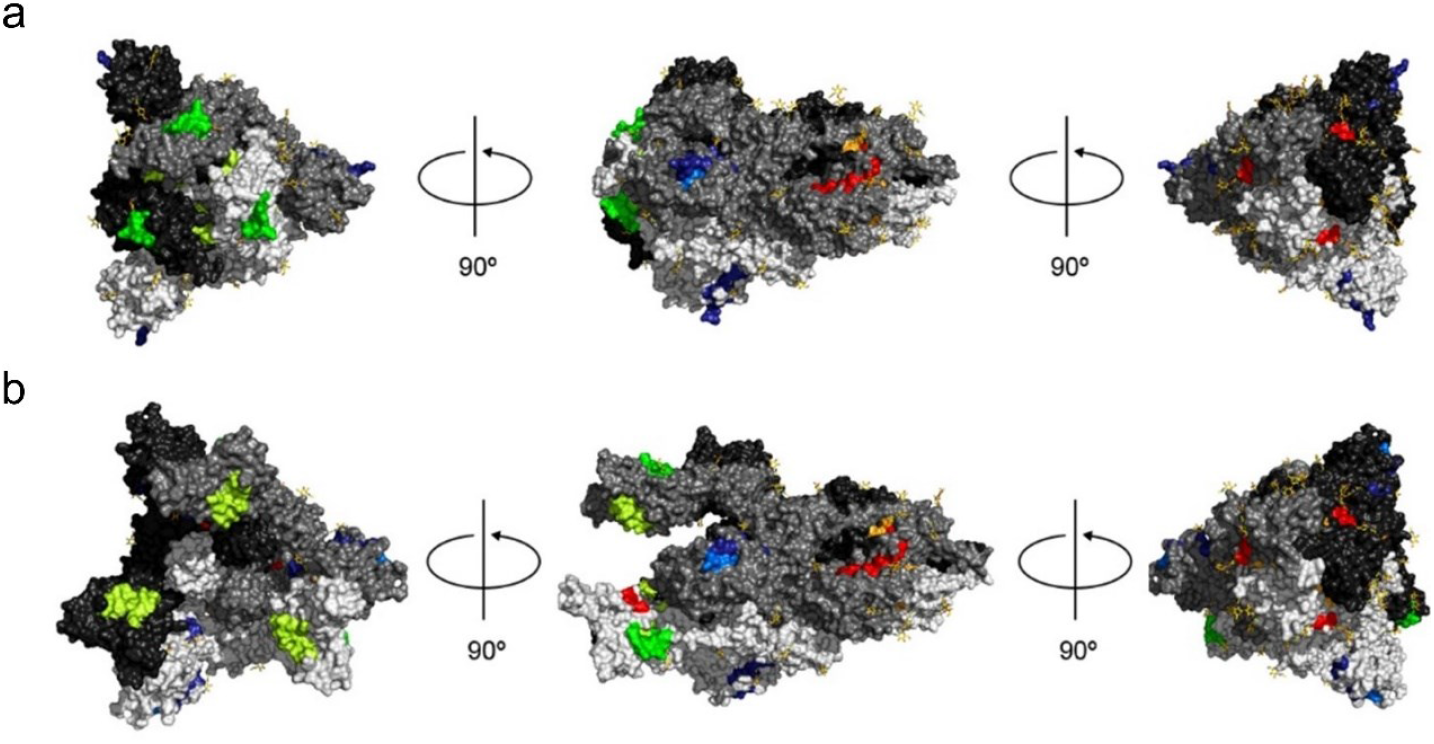
Peptide mapping and solvent exposure. **a** Experimental model of the Spike protein in a “closed” conformation (PDB code: 6VXX). The protein chains forming the trimer are displayed in different shades of grey. Carbohydrate moieties are shown in stick representation with carbon atoms in yellow. **b** Experimental model of the S-spike protein in “up” conformation (PDB code: 7KMS). This conformation of the S-spike protein allows for ACE2 binding. Peptides (P1-P8) are marked using a colour scheme indicated in Fig. 1. Partially overlapped P5 and P6 peptides are marked in red.

A fraction of the peptides’ residues is solvent-exposed, with residues forming the P2 and P7 peptides being the most buried (Fig. 5a, b). The P8 peptide is located in the C-terminal region of the protein that is disordered and therefore is most likely exposed to solvent. The residues corresponding to the selected peptides are solvent-exposed in both “closed” and “up” conformations^42^. To note, the change from “closed” to “up” conformation significantly increases solvent accessibility of the P4 peptide.

Another interesting feature of the selected peptides is that despite their relatively large distance in terms of protein sequence, some of them are quite close to each other in terms of their spatial location. For example, the P1 and P2 peptides are next to each other (Fig. 5a, b), and the P3 peptide and a fragment of the P4 peptide are in close vicinity in the closed conformation of the Spike. In addition, the P5 and P6 peptides (which partially overlap) together with the P7 peptide are also relatively close in the three-dimensional protein structure.

The analysis of the Spike protein (“up” conformation) in a complex with ACE2^43^ illustrates how the residues forming the P3 and P4 peptides, which correspond to the receptor binding domain (RBD) fragments, are still exposed to solvent (Fig. 6). This suggests that they can be parts of epitopes recognised by human antibodies, and they are not critical for the formation of the Spike protein-ACE2 interface (for modifications and properties of the peptides).

**Fig. 6.**
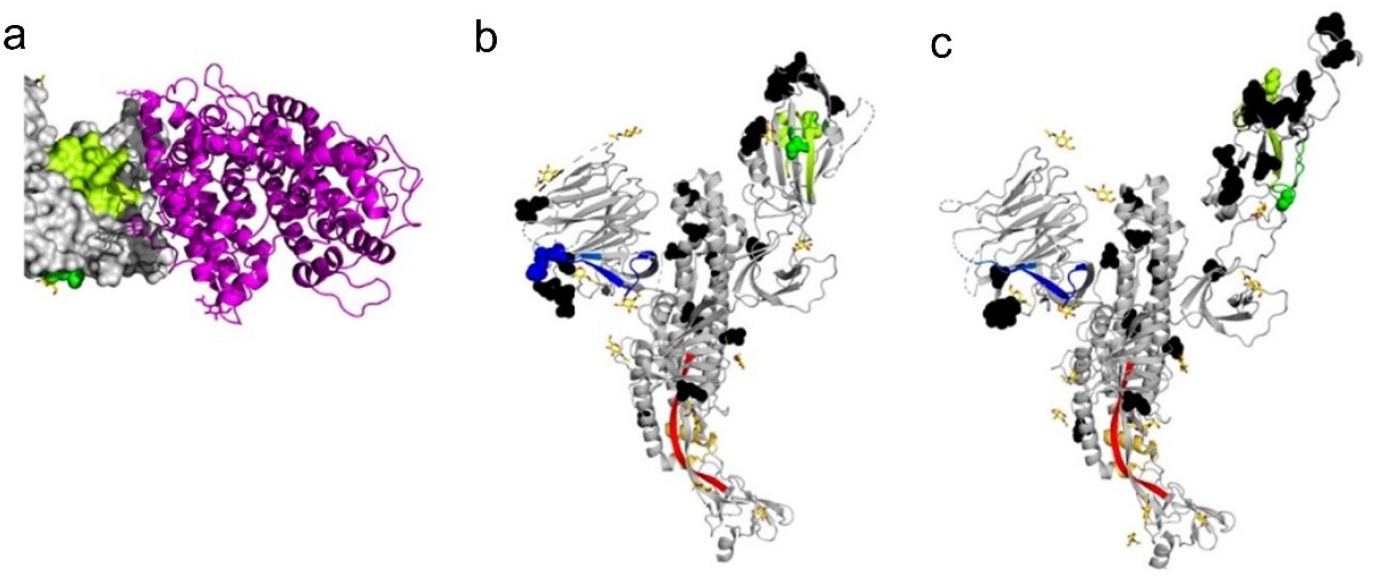
Impact of conformational changes, ACE2 binding and mutations on the selected peptides and in the Omicron/BA.1 variant. **a** Interaction between the RBD (shown in surface representation) and human ACE2 (in magenta; shown in ribbon representation). Residues forming the P3 and P4 peptides are displayed in green and lemon green, respectively. **b** Experimental model of the protein in “closed” conformation (PDB code: 6VXX). The protein chains forming the trimer are displayed in different shades of grey. Carbohydrate moieties are shown in stick representation with carbon atoms in yellow. **c** Experimental model of the S-spike protein in “up” conformation (PDB code: 7KMS). This conformation of the S-spike proteins allows the ACE2 binding. Identified peptides are marked using a colour scheme indicated in Fig. 1. Partially overlapped P5 and P6 peptides are marked in red. The PDB deposit (7KMS) corresponding to the S-spike-protein-ACE2 complex was used to generate this figure.

## 3. Discussion

An essential challenge in the development of preventive vaccines is to design an efficient and safe antigenic composition that ensures long-term stability and resistance against future mutations.

At the beginning of the COVID-19 pandemic (February-March 2020), we used our novel approach “Multi-Stable Epitope Sequencer” to identify a set of SARS-Cov-2 Spike regions (P1-P8), which were predicted to be stable against long-term future mutations. The World Health Organization defined the beginning of the pandemics in March 2020 and its end in May 2023. We have tested here all the viral lineages appeared within the period March 2020 to January 2024.

Our analyses confirm that the eight peptide sequences have been preserved in 97% of all 1,514 SARS-CoV-2 lineages reported in the CoV-GLUE dataset (the Pango Lineage Resources)^32^. Likewise, more than 90% of these peptides remained invariable at a global scale across the 49 predominant viral variants circulating throughout the pandemic (ECDC-WHO). In addition, these results have been confirmed in another analysis covering all non-synonymous mutations reported in the CoV-GLUE project for the all 28 variants considered of most interest (94.1%).

Lastly, the eight epitopes (P1-P8) were interrogated against the 3,362 complete viral genomes and 5,228,435 sequences, including mutations with low frequencies (0.0001) and the obtained results have shown that our epitopes are located in Spike protein cold spots (conserved regions with very low mutational frequency). Mutations occurring in the SARS-CoV-2 Spike do not follow a uniform distribution: some peptide sequences display high mutation rates while other small regions are remarkably conserved, and our predictive method was able to recognize these specific regions with high or low mutation frequency.

Our exhaustive analysis, spanning the whole pandemic period (March 2020 to January 2024), confirms the mutational stability of the selected peptides using our predictive approach capable of anticipating possible future viral variations.

We have confirmed that our “Multi-Stable Epitope Sequencer” technology is capable of reliably predicting stable epitopes against SARS-CoV-2 mutations that remain stable for relative long periods of time.

This work opens up a perspective for the development of advanced vaccines and therapies that target epitopes stable against future pathogen mutation and evolution. Designing preventive vaccines and neutralizing antibodies with antigens resilient to pathogen mutations is a pathway for the development of next-generation universal vaccines, valid for most or all future variants, and more efficient post-infection therapies.

## 5. Methods

### *5*.*1. Multi-Stable Epitope Sequencer*, methodology to design stable epitopes against future mutations. General procedure

Over the last 11 years, we have been performing exhaustive studies aiming to develop a new method to improve the traditional approaches in vaccine development^44^. This methodology allows the computational design-selection of specific epitopes with high probability to be stable against future mutations over a long period of evolution. The design and selection of these specific epitopes is a previous step for their synthesis and subsequent preclinical and clinical evaluation.

At the beginning of the COVID-19 pandemic, we studied the amino acid sequences of the first 22 variants of the S-spike entire protein deposited in the GISAID project database (February 2020). The objective was to consider the minimum number of amino acids that make up epitopes with a possible immunogenic response. These amino acid sequences should be recognized by the molecules of the major histocompatibility complex classes I and II. Specifically, we used a sliding window technique to extract all the possible sequences of length 9 and 15 amino-acids, and store the unmatched peptides. The final result of this analysis gave us a total of 3,112 possible epitopes. It has to be outlined that the goal of this procedure is not to predict the future from 22 early genomes, but to exclude likely variable regions and optimize the selection of stable, immunogenic peptides using a combinatorial search across all possible epitope combinations We first estimated the Class I immunogenicity (i.e., likelihood of T cell activation) of each epitope, and then separately assessed the peptide’s binding affinity to MHC molecules. For this, we used a bioinformatic tool developed by the IEDB team (http://www.iedb.org). This tool allows the epitopes to be classified according to their immunogenicity (Class I), which is estimated using their amino acid composition and order of amino acids^45^. We applied this tool to all the 9-mers (9 amino acid long sequences) and ranked them from highest values of such estimated immunogenicity to lowest. Since a negative estimation meant low probabilities of being immunogenic, we only considered the values for positive estimations and assigned a value of 0 to the rest of them.

Next, we evaluated the potential epitopes of length 9. In particular, we considered the reference IEDB allele, composed by 27 HLA molecules (A*01:01, A*02:01, A*02:03, A*02:06, A*03:01, A*11:01, A*23:01, A*24:02, A*26:01, A*30:01, A*30:02, A*31:01, A*32:01, A*33:01, A*68:01, A*68:02, B*07:02, B*08:01, B*15:01, B*35:01, B*40:01, B*44:02, B*44:03, B*51:01, B*53:01, B*57:01, B*58:01)^46^. Only those that obtained the best computational results were considered (i.e., those below the 1% cutoff on the percentile rank). Finally, we removed all the 9-mers which did not pass either the immunogenicity threshold, or the HLA-I binding threshold.

Then, we used the tool for Class II molecules^47^ to select the potential epitopes of length 15 amino-acids^47^. Likewise, we studied the affinity of each epitope to the most representative set of HLA alleles in the population (DRB1*03:01, DRB1*07:01, DRB1*15:01, DRB3*01:01, DRB3*02:02, DRB4*01:01, DRB5*01:01)^48^, and considered those that obtained the best computational results (i.e., those below the 10% cutoff on the percentile rank) as potential epitopes.

In order to develop a possible universal candidate, the epitopes obtained in the previous steps were weighted by the absolute frequency of the different HLA alleles that occur in the world population. Thus, our approach would favor the selection of peptides with higher affinity to the most prevalent histocompatibility molecules in the general population. This analysis was carried out considering experimental data (http://www.allelefrequencies.net/)^49^.

Once the list of potential epitopes was obtained it was necessary to select the most stable ones against future mutations. *This is the central aspect of our methodology*, which allows to obtain vaccine candidates with a higher probability of remaining stable over a determinate viral evolution.

To make such selection, we have relied on combinatorial optimization to develop the concept of “*λ superstring*” method (search procedure for a potential stable epitope against future mutations), proposed in a first scientific publication in 2015^44^. Later, we have *generalized* the notion of “*λ-superstrings*” to the weighted case for taking into account the predicted immune responses (2019)^50^. This generalization entails an important improvement in the applications to vaccine designs, as it allows epitopes to be weighted by their immunogenicities. More recently, we have developed the concept of “Multi-stable String”, a search procedure for a stable set of epitopes against future mutations. All these concepts are explained in detail below.

In summary, we used our methodology of combinatorial optimization to winnow the list of candidate sequences to identify possible immunologically active regions of the S-protein structure most likely to remain stable to future mutations. This analysis enabled selection of the eight short peptides (P1-P8) of the SARS-Cov-2 S-protein with the capacity to preserve high stability against variants of SARS-CoV-2.

### 5.2. “Multi-Stable String” concept

To obtain vaccine candidates consisting of a set of epitopes with elevated stability against long-term mutations require the solutions of the optimization problem to fulfill the λ superstring condition^44^, which is a part of the *Multi-Stable String* method. In particular, in a first approach, we seek to solve the *shortest λ superstring* problem (which generalizes the *shortest common superstring problem*, and the *set cover problem*, whose computational complexity is NP-hard). In short, we summarise the λ superstring condition and the *shortest λ superstring* problem as follows: Let *A* be a finite alphabet (in our case, formed by 20 amino acids) and 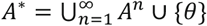 {θ} the set of all the possible strings formed by elements of *A*, where θ is the empty string. The set *A*^∗^ is a semigroup for the concatenation operation (denoted by +) where *t* + *t*′ = (*t*_1_, . . ., *t*_*n*_) + (*t*_1_′, . . ., *t*_*m*_′) = (*t*_1_, . . ., *t*_*n*_, *t*_1_′, . . ., *t*_*m*_′). We consider that *t* = (*t*_1_, . . ., *t*_*m*_) is a substring of another string *h* = (*h*_1_, . . ., *h*_*n*_) when ∃*k* ∈ {1, . . ., *n* − *m* + 1}| *t*_*k*+*i*−1_ = *h*_*i*_, ∀*i* ∈ {1, . . ., *m*}. Then, the overlapping between two strings *t* and *t*′ is defined as *overlapping*(*t, t*′) = max{*i* ∈ {0,1, …, *min*{*m, n*}} | *t*_*n*−*i*+*j*_ = *t*′_*j*_, *for j* = 1, …, *i*}. Additionally, if we consider *T* ⊆ *A*^∗^ the set of target strings (in our case, corresponding to potential epitopes), and λ ∈ ℕ, we can define a λ superstring as: let *H*_1_, …, *H*_*k*_ ⊆ *A*^∗^ and *T* ⊆ *A*^∗^, if we denote as *C*(*h, v*) the set of all common substrings of *h* and *v*, a λ superstring for the set (*H*_1_, …, *H*_*k*_, *T*) is a string *v* ∈ *A*^∗^| |*C*(*H*_*i*_, *v*) ∩ T| ≥ λ, ∀ *i* = 1, …, *k*.

Now we can enunciate the *shortest λ superstring* problem as: let *H*_1_, …, *H*_*n*_ ⊆ *A*^∗^ be a finite set of strings of the alphabet *A*, let *T* ⊆ *A*^∗^ the set of target strings, and let λ ∈ ℕ, find a *λ* superstring *v* ∈ *A*^∗^ for (*H*_1_, …, *H*_*k*_, *T*) with minimum length. This concept was generalized in^50^, where we included weights for each target string (epitopes). A weighted *λ* superstring for (*H*_1_, …, *H*_*k*_, *T, w*) is defined as a string *v* ∈ *A*^∗^ such as

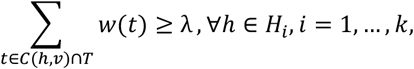

and solving the shortest weighted λ superstring problem is to find, for a given λ, a weighted λ superstring for (*H*_1_, …, *H*_*k*_, *T, w*) of minimum length.

In order to obtain a combination of short peptides to build a multi-peptide pool, we have adapted the problem of weighted λ superstring to the *Multi-stable string* problem, which is to find a group of short weighted λ superstrings obtained sequentially, each from a smaller subset. We first obtained a weighted λ superstring *v*_1_ for (*H*_1_, …, *H*_*k*_, *T*_1_, *w*); we next removed from *T*_1_ the target strings of lengths between 9 and 15 that were elements of *T*_1_, and also substrings contained in *v*_1_, obtaining the set *T*_2_; we subsequently obtained a weighted *λ* superstring *v*_2_ for (H_1_, …, *H*_*k*_, *T*_2_, *w*); this procedure was repeated until we obtained the group of strings as the final solution.

### 5.3. “Multi-stable string” solution

To solve the *Multi-stable string* problem, we first approached the *shortest common superstring problem* by developing an algorithm based on Estimation of Distribution Algorithms (EDA). This family of algorithms searches for the probability distribution of the best solution to a given problem with respect to an objective function (in our case, the λ parameter), starting with an initial distribution and evolving during the learning process, where the probability distribution is improved.

In short, the algorithm for solving the *shortest common superstring problem* goes as follows: Given a set of strings *s*_1_, …, *s*_n_ ⊆ *A*^∗^ over an alphabet *A*, we define the weight matrix as 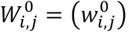, where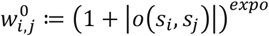. This matrix *W*^0^ will determine the initial estimation of the probability distribution of the *common superstring*, and the *expo* parameter is a control parameter of the algorithm.

Given a number of iterations *nit*, iterating, we then build the weight matrix *W*^k^ for k = 1, …, *nit*, which will let us estimate the probability distribution of the string of the *k* + 1 iteration. Given a population of size *spop*, we build the matrix *W*^k^ by sampling *spop* permutations and using the probability distribution given by *W*^k−1^, i.e., we generate π_1_, …, π_*s*pop_ permutations of the set {1, . ., *n*}.

The procedure in greater detail is as follows:

We determine a permutation π_i_ for each element of the population (spop times): first, we obtain a permutation π(1), from {1, . ., *n*} using the continuous uniform distribution, and we proceed iteratively:

1. If π(1), …, π(r) permutations (with *r* ≤ n − 1) have been chosen, we chose randomly a value for *b* ∈ {0,1}.
  a. If *b* = 0 (element on the left), we chose an element *u* ∈ *G* = {1, . ., *n*} − {π(1), …, π(r)} following the probability distribution *p* defined ∀*x* ∈ *G* as 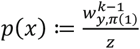, where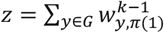. Using the value obtained for *u*, we redefine the set of permutations as (π(1), …, π(r + 1)) ≔ (*u*, π(1), …, π(r)), being π(1), …, π(r) the previous values.
  b. If *b* = 1 (element on the right), we chose an element *u* ∈ *G* = {1, . ., *n*} − {π(1), …, π(r)} following the probability distribution *p* defined ∀*x* ∈ *G* as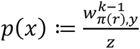, where 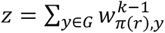 . Using the value obtained for *u*, we redefine the set of permutations as (π(1), …, π(r + 1)) ≔ (π(1), …, π(r), *u*), being π(1), …, π(r) the previous values.
2. If π(1), …, π(n − 1) permutations have been chosen and {*u*} = {1, . ., *n*} − {π(1), …, π(n − 1)}, then
  a. If 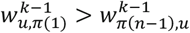, we redefine (π(1), …, π(n)) ≔ (*u*, π(1), …, π(n − 1)).
  b. Otherwise, (π(1), …, π(n)) ≔ (π(1), …, π(n − 1), *u*).

Once π_1_, …, π_*s*pop_ are obtained, we build the string 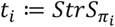 for each permutation

π_i_, merging (with overlap) the strings *s*_π(1)_, …, *s*_π(n)_, and we evaluate the length of *t*_*i*_. After fixing an acceptance *ratio*, we next chose a total of *m* = ⌊ratio · *s*pop⌋ permutations 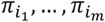 for which the lengths of 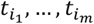 are the smallest ones.

Finally, in order to obtain *W*^k^, we begin by assigning *W*^k^ ≔ *W*^k−1^, and carry out the next readjustment for *j* = 1, …, *m* and for *l* = 1, …, *n* − 1:

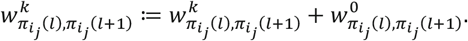

Thus, the factor (1 + |*o*(*s* _i_, *s*_j_)|)^*ex*po^ is implicitly considered. The reason for considering this factor is no division by 0 occurs. Additionally, it preserves the diversity in the population, allowing the appearance of two consecutive disjoint strings. Lastly, the output of the algorithm is the shortest *t*_*i*_ obtained after *nit* iterations.

The pseudocode of this algorithm is the following:

#### Algorithm 1

Algorithm for solving the *shortest common superstring* problem

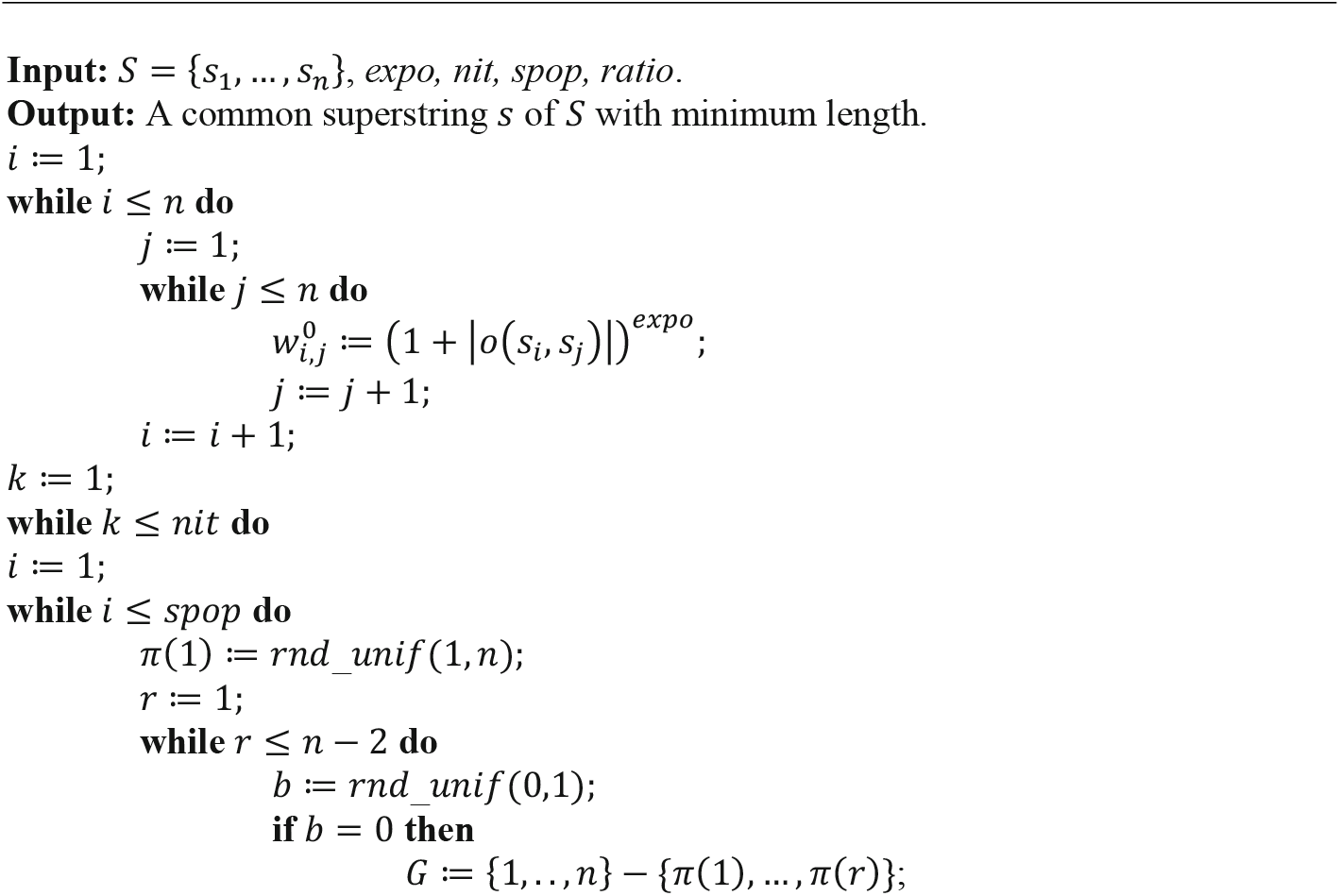
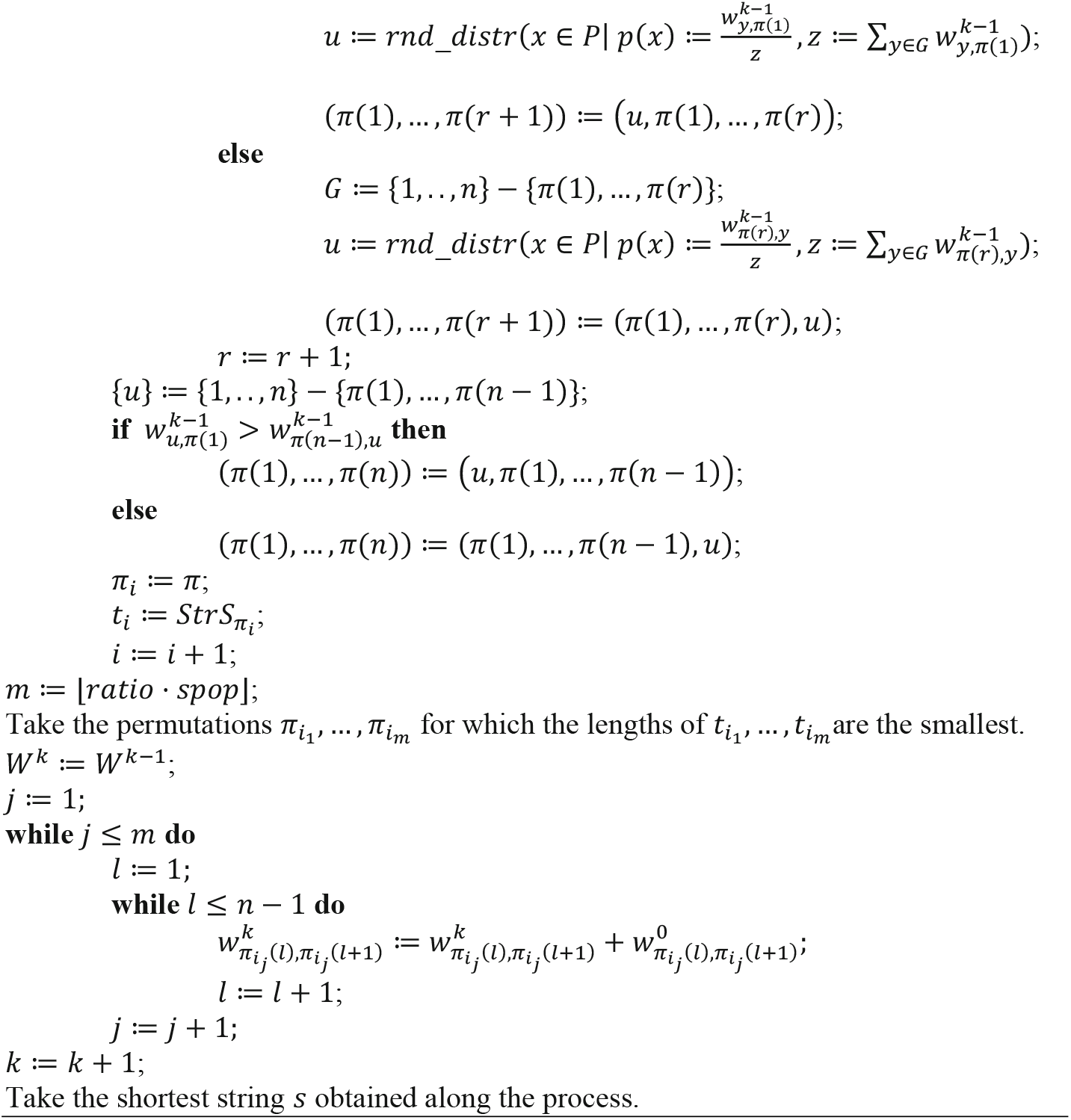

Finally, before applying the algorithm over the host string, target string, and weight sets, we adapted it, first, to solve the weighted λ superstring problem (requiring the combinatorial condition to our solutions; for more information, see^44,50^), and then, to solve the *Multi-stable string* problem (by applying the algorithm sequentially, and altering the target string set).

Therefore, given two sets of strings, a set of host strings, which models a set of instances of a protein (which in our case will be amino acid sequences of the protein for a given pathogen), and a set of target strings, which models a set of epitopes, a λ-superstring was defined to be a string that models a candidate vaccine containing, as substrings, at least λ target strings from each host string. This means that the vaccine covers at least λ epitopes in each patient. The associated optimization problem is to find a λ-superstring of minimum length, which means to find a candidate vaccine as short as possible.

### 5.4. Analysis of the ten highly conserved antigenic sequences not selected (R1-R10)

Our methodological system selected eight short SARS-CoV-2 spike peptides (P1-P8) following inspection of millions of possible amino acid combinations, but it evaluated and discarded those that were unlikely to interact with HLA. In particular, it evaluated and discarded the aforementioned ten highly conserved antigenic sequences. This second set of 10 S-spike peptides (R1-R10) was detected but not selected due to the following criteria:

First, based on the 22 S-spike sequences considered (February-March 2020), another main element of our program was maximised to obtain the best epitopes, in addition to fulfilling the lambda-superstring criterion. This point corresponds to the estimation of the Class-I immunogenicity by the tool “T cell Class-I pMHC immunogenicity predictor” (http://tools.iedb.org/immunogenicity)^51^, which classifies epitopes according to their immunogenic response, amino acid composition, and order of their amino acids sequence^45^. After estimating the values of the “second set of ten stable epitopes”, we observed that seven of them scored less than the smallest score obtained by our main set of eight selected stable epitopes (which scored higher than 0.153). Consequently, since a higher value is related to a higher probability of generating an immune response, those seven were outperformed by the chosen ones because of their lower estimated immunogenicity.

Therefore, only three of the “second set of ten stable epitopes (R1-R10)” were possibly immunogenically good candidates. Two of them (the sequences NITNLCPFGEVFN and KLNDLCFTNVYADSFVIRGDEV) overlapped with two of our eight stable epitopes (P3 and P4), and are discussed below. NITNLCPFGEVFN of length 13, overlapped with our selected sequence P3 (GEVFNATRFA) of length ten. This sequence was not considered because, despite of being longer than P3, the estimated immunogenic response of sequence P3 scored 0.312, while the sequence belonging to the “second set of ten stable epitopes” only scored 0.276.

In addition to considering the score of the tool described above, we also took into account the affinity of short peptides for Class I molecules^52^ (http://tools.iedb.org/mhci)^53^ and the affinity of long peptides to Class II molecules^47^ (http://tools.iedb.org/mhcii)^54^. Additionally, each MHC molecule was weighted by the absolute frequency of the different HLA alleles that occur in the population to develop an antigenic set as universal as possible (http://www.allelefrequencies.net)^49^. Consequently, we maximized a combined score when the values obtained with these tools were taken into account.

The 22-residue epitope KLNDLCFTNVYADSFVIRGDEV that overlapped with our identified 24 residue P4 epitope (VYADSFVIRGDEVRQIAPGQTGK) was close in estimated immunogenic response (0.481 against 0.471 of the P4 peptide). However, when the combined score was taken into account, the former scored 7.136, while peptide P4 exhibited 7.778. This indicates that, if both estimated immunogenicity and binding affinities were taken into account, P4 peptide outperformed the epitope of length 22.

Next, the last sequence belonging to the set of ten stable epitopes (R1-R10), namely the 23-residue peptide RLDKVEAEVQIDRLITGRLQSLQ was compared with the identified P7 peptide (ALQIPFAMQMAYRFNGIGVTQNVL). They exhibited similar lengths and estimated immunogenicity. When the estimated immunogenicity was calculated, the first scored 0.1666, and the second 1.586. However, the differences when considering the binding affinity in the combined score were much higher, yielding values of 6.289 for the 23-length epitope, and 8.038 for the P7 peptide.

Finally, when we compared the two epitope pools (P1-P8 and R1-R10), both the estimated immunogenicity and the combined score achieved by our main set of eight stable P1-P8 epitopes (0.418±0.344, mean±std; and 4.581±3.8, respectively) were higher than the scores calculated for R1-R10 (-0.097±0.415; and 2.847±2.381, respectively).

### 5.5. Peptide analysis

The Spike protein sequence that was used in our analysis was obtained from Uniprot (code: P0DTC2; SPIKE_SARS2), and is shown below. The sequences corresponding to our identified peptides are highlighted.

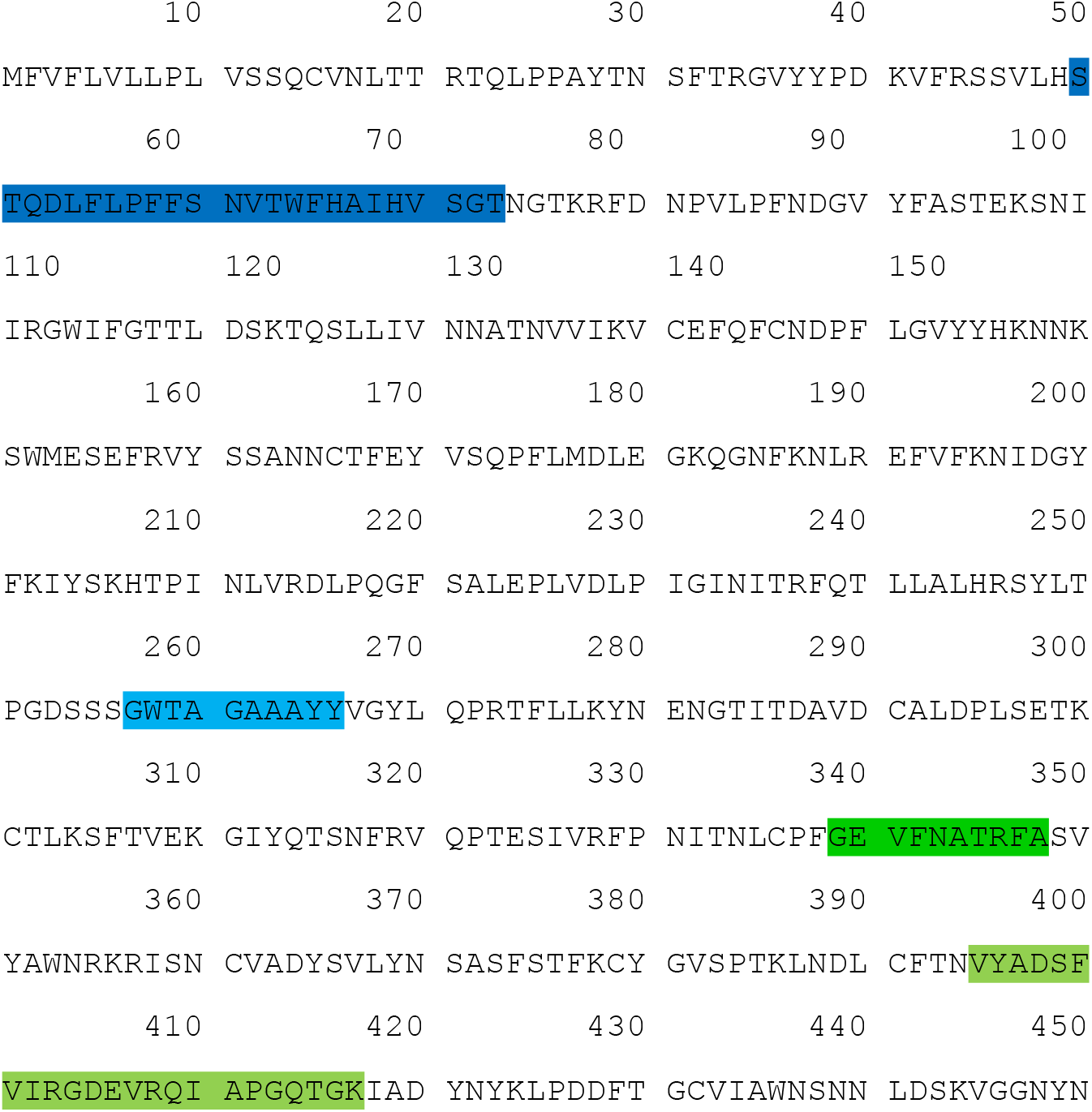
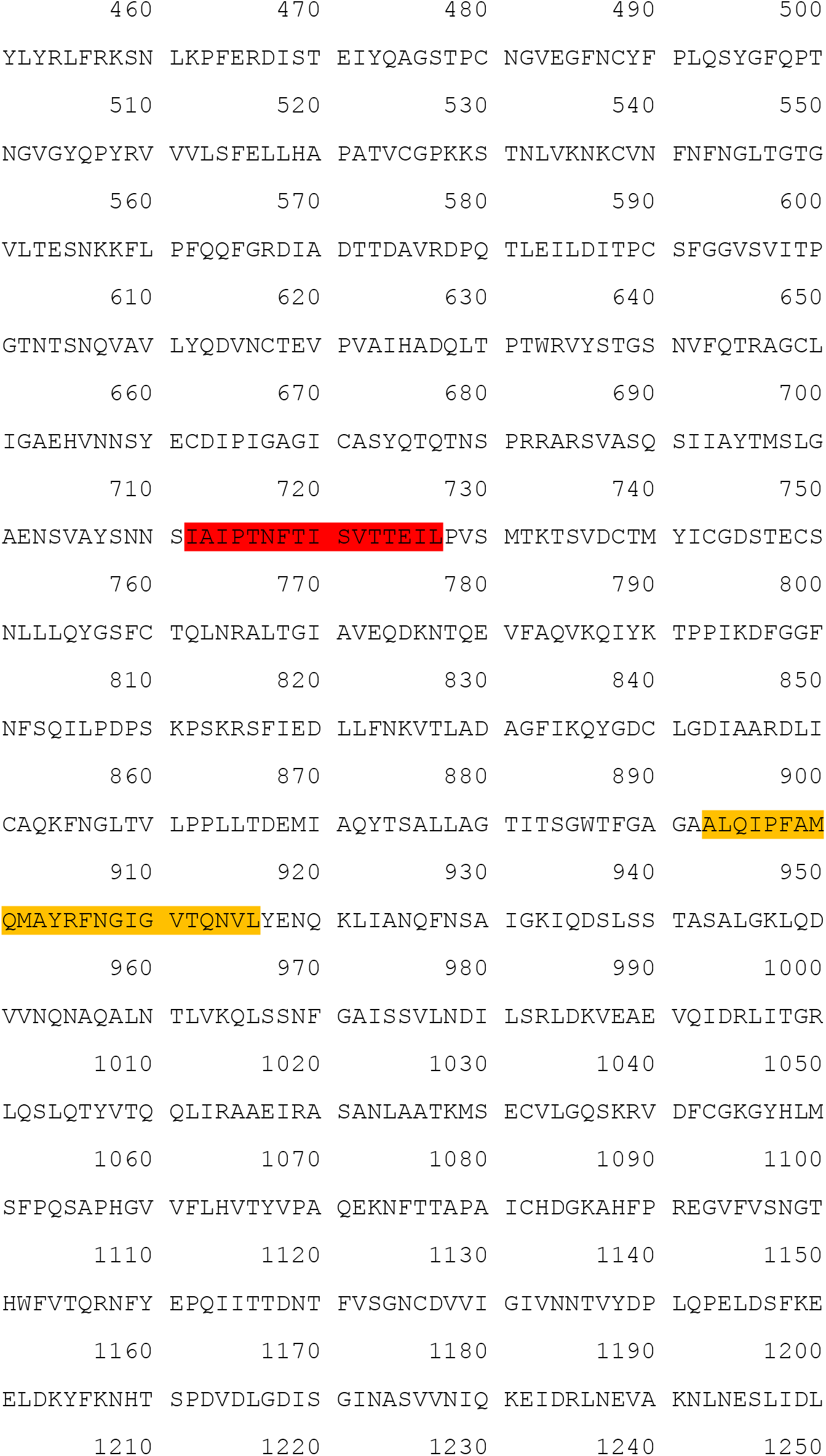
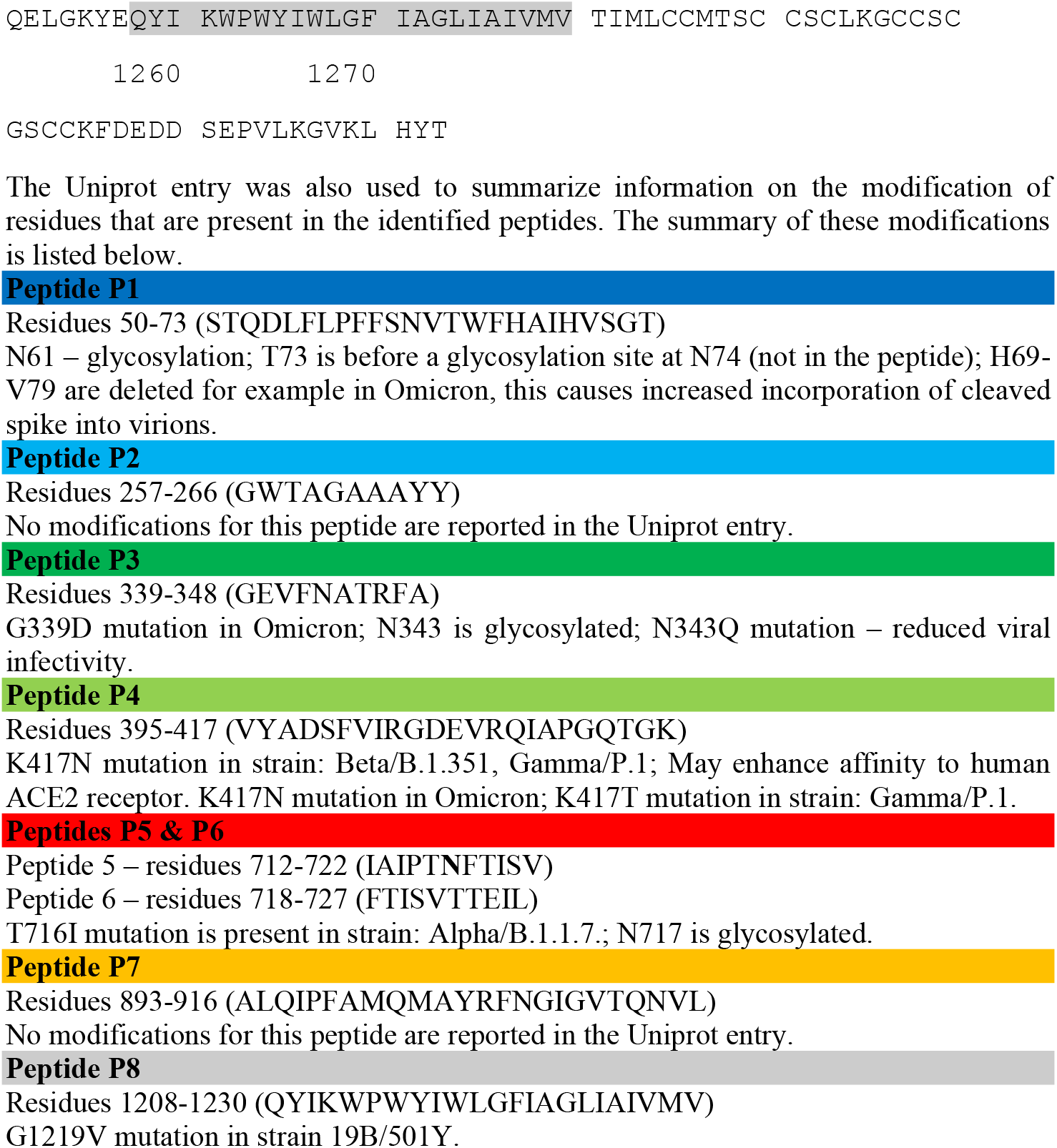

### 5.6. Sequence variation analysis (Results, section #2.5)

We analyzed publicly available variation data from GISAID^36^ to test the stability of the epitopes against mutations. These include 3,854 complete genomes, across 1,514 SARS-CoV-2 lineages (available at Nextstrain^34^) from which the entropy per site and the number of mutational events across the strain tree are computed (pandemic period March 2020 to January 2024). Likewise, the protein variation analysis of 5,228,435 GISAID hCoV-19 sequences, available at CoV-GLUE database^35^ has been taken into account in our study. Data from CovGlue was downloaded for all SARS-CoV-2 variants separately and disregarded mutations with a frequency lower than 0.0001. To study whether each residue had the same mutation probability, we performed a goodness of fit test of the distribution of probabilities using χ^2^ test scores based on values obtained from the number of non-synonymous mutation events (see Fig. 4c). We obtained a *p* − *valu*e = 0 and a χ^2^ − *s*tati*s*ti*c* = 2 × 10^4^, rejecting the null hypothesis. This indicates the probability of mutation is not the same in every region of the protein. The non-synonymous mutations are not uniformly distributed over the SARS-Cov-2 S-protein, and consequently, we can observe regions with high or low mutation frequency. The probability for a peptide of being invariable (e.g. identical to the reference protein sequence) in a given SARS-CoV-2 variant was approximated by assuming independence of the sites and multiplying the expected non-mutated frequencies (1 - mutation frequency). We compared these results with 10,000 random samples of seven non-overlapping peptides of the same size as the ones selected by our algorithm (note that 5 & 6 peptides overlap and are considered as a single peptide spanning 712-727 in this analysis). The selected epitope pool presented a frequency of 0.0023 mutations per site, and relative entropy of 0.022. This P1-P8 epitope pool is expected to be invariable in 71.4% of the sequences, as compared with 45.0% in 10,000 randomly sampled combinations of similar fragments from the Spike protein. The results indicate that P1-P8 epitopes are not located in mutational hotspots of the S-protein. The amino acid composition for these combinations of peptides was computed in terms of amino acid frequencies (%) and compared to the selected epitopes. Significant deviations were considered when observed values in the selected epitope set were higher or lower than the 95% and 5% percentile values in the distribution obtained for the 10,000 random peptide samples.

## Data Availability

Data have been made publicly available via the Global Initiative on Sharing Avian Influenza Data (GISAID) database, GISAID-Outbreak project, CoV-GLUE database, Nextstrain project and The Allele Frequency Net Database (AFND).

## CRediT authorship contribution statement

**Ildefonso M. De la Fuente**: Conceptualization, Writing – original draft, Formal Analysis, Supervision, Methodology, Project Administration. **Iker Malaina**: Writing – review & editing, Formal Analysis, Software, Methodology, Funding Acquisition. **Maria Fedetz**: Writing – review & editing, Investigation, Data curation, Methodology, Supervision. **Maksymilian Chruszcz:** Investigation, Writing – review & editing, Formal Analysis. **Gontzal Grandes**: Investigation, Writing – review & editing, Formal Analysis. **Oleg Targoni:** Investigation, Writing – review & editing. **Antonio Abel Lozano-Pérez:** Investigation, Writing – review & editing, Funding Acquisition. **Eyal Shteyer:** Investigation, Writing – review & editing. **Ami Ben Ya’acov:** Investigation, Writing – review & editing. **Agustín Gómez de la Cámara:** Investigation, Writing – review & editing. **Alberto M. Borobia**: Investigation, Writing – review & editing. **Jose Carrasco-Pujante:** Investigation, Writing – review & editing, Visualization. **Jose Ignacio Pijoan**: Investigation, Writing – review & editing. **Carlos Bringas:** Investigation, Writing – review & editing. **Gorka Pérez-Yarza:** Investigation, Writing – review & editing. **Alberto Ouro:** Investigation, Writing – review & editing. **Ivan Sanchez:** Investigation, Writing – review & editing. **Jesus M. Cortes**: Investigation, Writing – review & editing. **Michael J. Crawford:** Investigation, Writing – review & editing. **Varda Shoshan-Barmatz:** Investigation, Writing – review & editing. **Vladimir Zhurov**: Investigation, Writing – review & editing, Formal Analysis, Visualization. **José I. López**: Investigation, Writing – review & editing, Formal Analysis. **Shira Knafo:** Investigation, Writing – review & editing. **Magdalena Tary-Lehmann**: Investigation, Writing – review & editing. **Toni Gabaldón**: Investigation, Formal Analysis, Writing – review & editing. **Miodrag Grbic**: Conceptualization, Investigation, Writing – review & editing, Formal Analysis, Supervision, Funding Acquisition.

## Acknowledgments

We would like to thank Andrea O’Malley for reading the manuscript and valuable comments. M.G. acknowledges the epidemiology input of M. Gotovac. Dr. Lozano-Pérez acknowledges the European Commission ERDF/FEDER Operational Program ‘Murcia’ CCI Nº 2007ES161PO001 (Project No. 14-20/20). M.G acknowledges support from the NSERC Discovery grant (Canada). This work also was supported by the Department of Education of the Basque Government via the Consolidated Research Group MATH MODE (IT1456-22). Furthermore, IMDF and IM were supported by the UPV/EHU and Basque Center of Applied Mathematics, Grant US21/27. The funders of the study had no role in the study design, data collection, data analysis, data interpretation, writing of the report or the decision to submit it for publication.

